# Evidence of Resiliency in Maternal Health Services and Outcomes in Kono District, Sierra Leone during the COVID-19 Pandemic: An Observational Study

**DOI:** 10.1101/2025.01.22.25320951

**Authors:** Foday Boima, Zeleke Abebaw Mekonnen, Manso M Koroma, Marta Lado, Adrienne K. Chan, Sharmistha Mishra, Alhaji U. N’jai, Bailah Molleh, Stephen Sevalie, Sulaiman Lakoh

## Abstract

**Background:** This study evaluated the resilience and outcomes of maternal health services during the COVID-19 pandemic in two health facilities in eastern Sierra Leone. Its aims to describe the use of maternal healthcare services and maternal and neonatal outcomes in these two facilities before, during, and after the COVID-19 pandemic.

**Methods:** The study involved analysis of routine programme data (March 2019 to February 2022) from two public-funded health facilities supported by a non-governmental organization (Partners In Health in Sierra Leone): Koidu Government Hospital and Wellbody Clinic. Aggregated and de-identified secondary data from the Partner In Health Maternal Health Database was abstracted using a standardized tool. Descriptive statistics and bivariable negative binomial regression were used to assess the association between time periods (before COVID-19 [March 2019 to February 2020], during COVID-19 emergency [March 2020 to February 2021], after COVID-19 emergency [March 2021 to February 2022) and outcomes each month (fourth antenatal care visit and facility deliveries).

**Results:** The study analyzed 3,204 fourth antenatal care visits and 7,369 deliveries over 36 months at both health facilities. Fourth antenatal care visits (from 947 to 920) and facility deliveries (from 2309 to 2221) decreased during COVID-19 compared to pre-COVID-19. However, maternal (from 32 to 23) and neonatal (36 to 26) deaths declined during COVID-19 compared to the pre-COVID-19 period at Koidu Government Hospital.

**Conclusion:** In Sierra Leone, the resources and efforts directed to the post-Ebola recovery strategy were tested during and after the COVID-19 pandemic. Our study demonstrates the resilience of maternal and neonatal services in two healthcare facilities in a less-affected region of Sierra Leone, to the anticipated disruptions due to the COVID-19 pandemic.

## Introduction

Sierra Leone has one of the highest maternal mortality rates in the world (1). For every 100,000 live births, an estimated 717 women die from causes related to pregnancy and childbirth in the country (1). The neonatal mortality rate in Sierra Leone is 31 deaths per 1000 live births and remains far from the Sustainable Development Goal target of less than 12 per 1000 live births by 2030 (2). Key to reducing maternal and neonatal mortality is an effective health system platform to provide maternal health services, including deliveries and operative care (such as caesarean sections) (2). Services provided within health facilities are vulnerable to structural shocks from infectious disease epidemics. Epidemics can lead to a shortage in the health workforce caused by healthcare-associated infections and fear among patients of acquiring infection in health facilities (3). Public health emergencies also lead to the redirection of clinical care from routine to epidemic-related services (4).

The 2014-2016 Ebola epidemic in Sierra Leone, with an estimated 14,000 confirmed cases and 3,965 deaths (5), led to large disruptions across the health system. The Ebola epidemic resulted in a 22% decline in antenatal care and delivery services at primary health units in Siera Leone (6), and a 20% decline in the number of hospital deliveries and caesarean sections (7), a pattern that was similar across all three West African countries severely affected by the Ebola outbreak (8–10). However, there was heterogeneity in the magnitude of disruptions to antenatal and obstetric care across districts in Sierra Leone (7). Districts with low numbers of confirmed Ebola cases experienced smaller reductions in antenatal care and facility deliveries (7).

Following the Ebola outbreak, the reproductive, maternal, neonatal, and child health programs remained a major priority for the Ministry of Health in Sierra Leone. The country’s health-system recovery agenda included training of healthcare workers, robust data management systems in facilities, and drug supply (11). These health-system recovery efforts were underway when the COVID-19 pandemic hit in early 2020. An example of the post-Ebola and pre-COVID efforts to improve maternal and neonatal services in health facilities was the partnership between the Ministry of Health and Partners In Health to support a primary and a secondary health facility in Kono district, in Eastern Sierra Leone. In the primary health facility (Wellbody Clinic), investments included mentorship for maternity staff and the establishment of maternal waiting homes where women with high-risk pregnancies receive 24-hour monitoring before their due dates. In the only secondary health facility in Kono District (Koidu Government Hospital), post-Ebola health-system strengthening included resourcing critical infrastructure resources: installing 24-hour electricity and running water, local production of oxygen supplementation, a fully operational blood bank, and a special care baby unit. Koidu Government Hospital is the only facility providing specialist care for pregnancy complications, including caesarean sections in Kono District(12).

There is some evidence at the national level, (8,13) but this study will narrow the focus to one district with 2 hospitals national data suggest disruptions in maternal health services, with a decline in first antenatal care visits and facility-based deliveries in Sierra Leone (8). However, Kono experienced a smaller, cumulative confirmed cases of 112 than the more urban regions in the country with national cumulative confirmed cases of 7681 COVID-19 outbreak (14). The post-Ebola recovery investments in Wellbody Clinic and Koidu Government Hospital provide an opportunity to conduct a more focused examination of facility-level maternal health services and maternal and neonatal outcomes in the context of the COVID-19 pandemic. We therefore sought to describe the use of maternal healthcare services and maternal and neonatal outcomes in these two facilities over time, before, during, and after the post-COVID-19 emergency in Sierra Leone.

## Methods

### Study Design

We applied a retrospective cross-sectional study design using facility-level programme data, to compare service use and health outcomes across three time periods; pre-, during, and post-COVID-19 pandemic. We utilized the STROBE (Strengthening the Reporting of Observational Studies in Epidemiology) guidelines in our study to ensure comprehensive and transparent reporting.

### Study Settings

Sierra Leone has a population of 7.5 million people (12) with three levels of service delivery provided across 1,468 healthcare facilities as of December 2023: peripheral health units (primary care), district hospitals (secondary care), and tertiary hospitals (tertiary care). This study was conducted in two Partners In Health-supported facilities, a non-governmental organization that works in partnership with the Sierra Leone Ministry of Health, in Kono district located in the eastern region with an estimated population of 600,000, of which 75% reside in rural areas (12): Wellbody Clinic (a primary health care facility, serving a population of 21,000) and Koidu Government Hospital (a secondary health care facility, serving a population of approximately 506,000 people) (**Figure 1**). Wellbody Clinic (with an average of 36 deliveries per month) provides basic emergency obstetric and newborn care. Koidu Government Hospital (with an average of 190 deliveries per month) provides comprehensive emergency obstetric care, including caesarian sections and onsite blood transfusion services.

**Figure 1.**
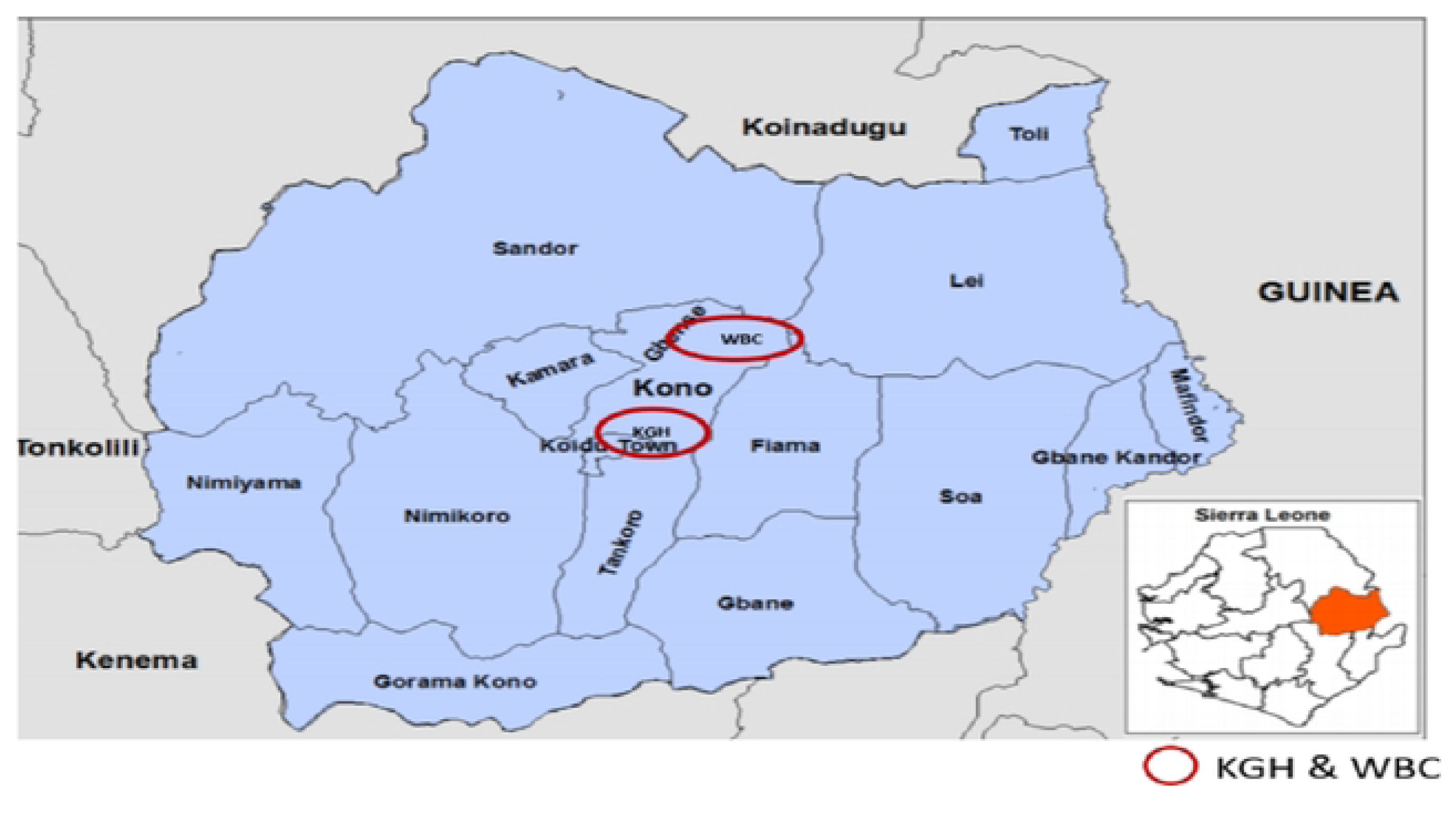
Map of the study area

### Study Population

*We reviewed the records* of pregnant and peripartum women and neonates who sought care at Wellbody Clinic and Koidu Government Hospital between March 2019 and February 2022.

### Data Sources

Data for this study was extracted from the Partners In Health Maternal Health Database on June 10, 2024. The database consists of secondary, de-identified, aggregate-level programme data reported monthly from the two health facilities which are aligned with nationally developed reportable data elements and indicators by the Ministry of Health.

### Variables

Our outcomes of interest were the monthly number of fourth ANC visits (ANC4), number of institutional deliveries, maternal mortality ratio (maternal deaths per 100,000 live births), and neonatal mortality ratio (neonatal deaths per 1,000 live births). These outcomes were selected because they represent key performance indicators for the healthcare system in Sierra Leone. We defined the following 12-month time periods of interest as follows: pre-COVID-19: (March 2019 to February 2020), during the COVID-19 emergency: (March 2020 to February 2021), and post-COVID-19 emergency: (March 2021 to February 2022). The dates were chosen because the first diagnosed case of SARS-CoV-2 in Sierra Leone was on March 31, 2020 (15).

### Data preparation and analyses

Monthly data on numerators for the outcomes listed above and relevant denominators (number of live births) were manually extracted from the Partners in Health Maternal Health Database and entered into an Excel database. The data in Excel was transposed into a long format from a wide format. Analyses were conducted in STATA15 (StataCorp LLC, College Station, TX), and included checks for missing data, data cleaning, and variable creation for analyses.

We used descriptive statistics to summarize outcomes by period. To address objective 1, we first visualized data on outcomes to describe trends in ANC4 visits and deliveries over the full study period, with demarcations to depict each period of interest. Next, to quantify the association between the period (exposure variable) and each count outcome (ANC4 visit; institutional delivery), we conducted bivariable negative binomial regression analysis (due to the observed over-dispersion for both count outcome variables)(16). We reported the incidence rate ratio (IRR) for each outcome with the corresponding 95% confidence interval (CI) using the during-COVID emergency period as the reference group. We interpreted observed differences between periods, based on the CI of the IRR. For objectives 2 and 3, we computed and described the maternal and neonatal mortality ratios for the three time periods, respectively. We did not conduct analyses of association due to the small number of outcomes.

### Ethics

Ethical approval was obtained from the Sierra Leone Ethics and Scientific Review Committee with approval number 018/05/2023. Routine secondary data was abstracted under a waiver of informed consent from the ethics committee and approval from the health facility management.

## Results

The study analyzed 3,204 fourth antenatal care visits and 7,369 deliveries over 36 months at both health facilities. There was a similar number of ANC4 visits pre-COVID-19 (947) and during the COVID-19 emergency (920), with a large increase post-emergency (1337), (**Table 1, Figure 2**) – a pattern that was observed in both study sites. A similar overall pattern was observed with institutional deliveries: from 2309 pre-COVID-19, 2221 during the emergency, and 2839 post-emergency (**Table 1, Figure 3**). There were no maternal deaths or neonatal deaths at Wellbody Clinic.

**Figure 2.**
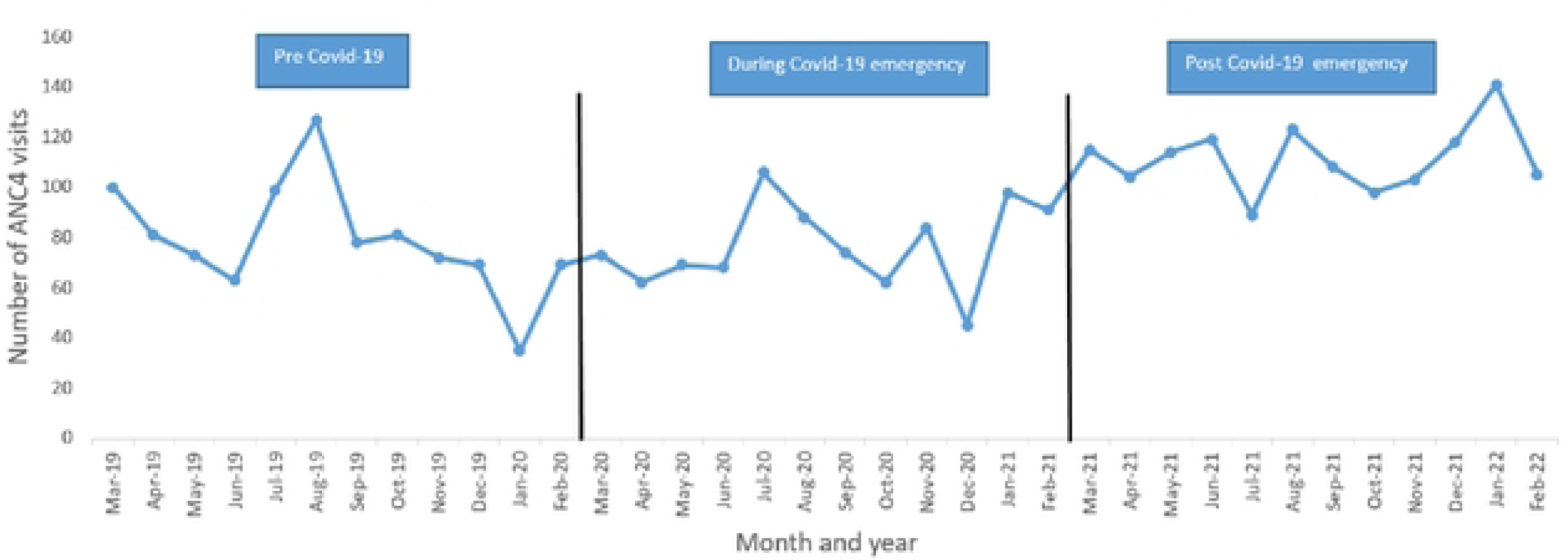
Monthly number of ANC4 visits at two health facilitiesIn Kono District, Sierra Leone, Man:h 2019-Feb 2022. ^0^ANC4 refers to the fourth antenatal clinic visit. *Pre-COVID-19 (March 2019-Feb 2020), duringCOVID-19 Emergency (March 2020-Feb 2021),Post COVID-19 Emergency (March 2021-Feb 2022).

**Figure 3.**
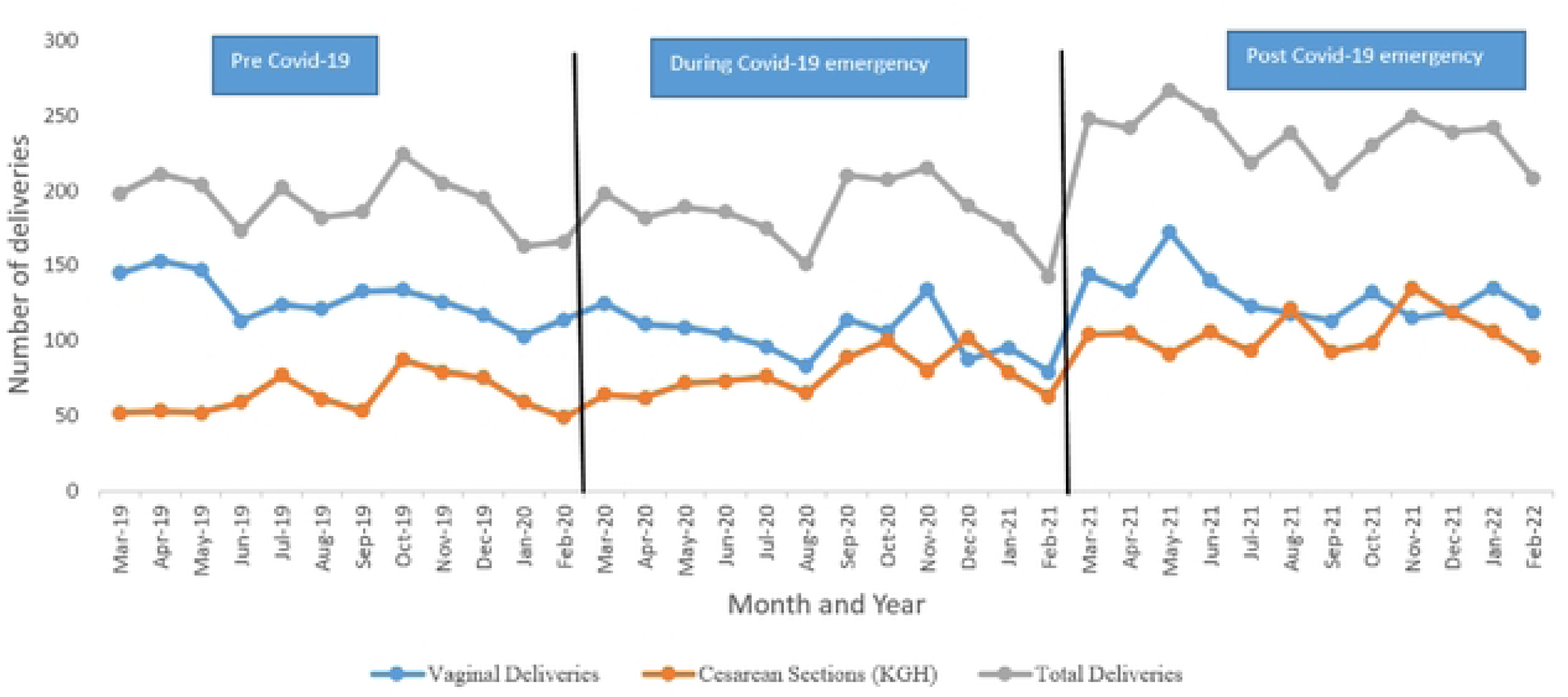
Monthly number of deliveries at two health facilities in Kono District, Sierra Leone, March 2019-Feb 2022. •Pre-COVID-19 (March 2019-Feb 2020), duringCOVID-19 Emergency (March 2020-Feb 2021),Post COVID-19 Emergency (March 2021-Feb 2022). **•KGH refers toKoidu Government Hospital.**

**Table 1:** Maternal health service use and outcomes in a primary and secondary health facility in Kono District, Sierra Leone before during, and after the COVID-19 emergency.

| Outcome | Number |  |  |
| --- | --- | --- | --- |
|  | Pre-COVID-19, March 2019 to February 2020 | During-COVID-19 emergency, March 2020 to February 2021 | Post-COVID-19 emergency, March 2021 to February 2022 |
| **ANC4 Visit** |  |  |  |
| Total | 947 | 920 | 1337 |
| Wellbody | 164 | 99 | 274 |
| KGH | 783 | 821 | 1063 |
| **Institutional deliveries** |  |  |  |
| Total | 2309 | 2221 | 2839 |
| Wellbody | 482 | 413 | 411 |
| KGH | 1827 | 1808 | 2428 |
| **Live births** |  |  |  |
| Total | 2246 | 2186 | 2785 |
| Wellbody | 488 | 415 | 414 |
| KGH | 1758 | 1771 | 2371 |
| **Maternal death** |  |  |  |
| Total | 32 | 23 | 25 |
| Wellbody | 0 | 0 | 0 |

| KGH | 32 | 23 | 25 |
| --- | --- | --- | --- |
| **Neonatal death** |  |  |  |
| **Total** | **36** | **26** | **17** |
| **Wellbody** | **0** | **0** | **0** |
| **KGH** | **36** | **26** | **17** |

\* KGH refers to Koidu Government Hospital, ANC4 refers to the fourth antenatal care visit
\* Pre-covid-19 (March 2019-Feb 2020), During covid-19 Emergency (March 2020-Feb 2021), Post covid-19 Emergency (March 2021-Feb 2022)

The monthly number of ANC4 visits fluctuated during each period, but the observed pattern suggests a steady increase over time and into the post-emergency period (**Figure 2**). Of note, there was a sharp decline in January 2020, two months before the first case of SARS-CoV-2 was detected in Sierra Leone, followed by rapid recovery in February 2020; and a similar pattern was noted in December 2020.

The monthly number of institutional deliveries demonstrated a similar pattern of a steady increase over the three time periods (**Figure 3**), albeit with fewer fluctuations than ANC4 visits. Temporary declines in deliveries occurred in January 2020, August 2020, and February 2021 – each followed by recovery to higher levels than just before each decline (**Figure 3**). With respect to the mode of delivery, the monthly number of vaginal deliveries appeared to temporarily decline during the COVID-19 emergency, whereas caesarian sections steadily increased over the three time periods (**Figure 3**).

### Impact of COVID-19 on maternal service utilization

Relative to the period of COVID-19 emergency, there were no observable differences in the rate of ANC4 visits during the pre-COVID-19 period [IRR=1.02, 95%CI: 0.61, 1.72] and during the post-COVID-19 emergency [IRR=1.45, 95%CI: 0.87, 2.42] (**Table 2**). Relative to the COVID-19 emergency period, there was also no difference observed in maternal deliveries: pre-COVID-19 [IRR=1.03, 95%CI: 0.69, 1.56] and post-COVID-19 emergency [IRR=1.28, 95%CI: 0.85, 1.92] (**Table 2**).

**Table 2.** Comparison in monthly ANC4 visits and institutional deliveries across three time-periods, in two facilities in Kono District, Sierra Leone, March 2019-Feb 2022.

| Time period | ANC4 visit |  | Total deliveries |  |
| --- | --- | --- | --- | --- |
|  | IRR | 95% CI | IRR | 95% CI |
| Pre Covid-19 | 1.02 | 0.61, 1.72 | 1.03 | 0.69, 1.56 |
| During Covid-19 Emergency | Reference |  | Reference (1) |  |
| Post Covid-19 Emergency | 1.45 | 0.87, 2.42 | 1.28 | 0.85, 1.92 |

\* IRR- Incidence rate ratio, CI- Confidence Interval, ANC- Antenatal care
\* Pre-covid-19 (March 2019-Feb 2020), During covid-19 Emergency (March 2020-Feb 2021), Post covid-19 Emergency (March 2021-Feb 2022)

### Impact of COVID-19 on maternal and neonatal outcomes

The maternal mortality ratio (per 100,000 live births) decreased over three time periods; 1,424 pre-COVID-19, 1,052 during the COVID-19 emergency, and 897 post-COVID-19 emergency. Similarly, the neonatal death ratio per 1,000 live births reduced from 16.0 pre-COVID-19, 11.9 during the COVID-19 emergency, and 6.1 post-COVID-19 emergency (**Table 3**).

**Table 3.** Maternal and neonatal outcomes across three time periods, in two facilities in Kono District, Sierra Leone, March 2019-Feb 2022.

| Outcomes | Pre-COVID-19<br>(March 2019-Feb 2020) | During COVID-19 Emergency<br>(March 2020-Feb 2021) | Post COVID-19 Emergency<br>(March 2021-Feb 2022) |
| --- | --- | --- | --- |
| Maternal Mortality Ratio<br>per 100,000 live births | 1,424 | 1,052 | 897 |
| Infant Death Rate per 1,000<br>live births | 16.0 | 11.9 | 6.1 |

## Discussion

Using routine program data, we found maternal services and outcomes remained relatively stable in two health facilities in Sierra Leone, a primary and a secondary care center. These findings differ from what the national-level trends had suggested (8). Except for the short-term reduction in ANC4 visits in early 2020 shortly before the first reported cases of COVID-19 in Sierra Leone, there was a stable l pattern in the uptake of maternal services and outcomes. Caesarian sections performed in the two facilities gradually increased into the post-emergency period. Although we were not able to conduct an analytic comparison due to small numbers, the overall maternal mortality ratio and neonatal mortality ratio declined over time – with the neonatal mortality ratio falling to well below national levels and the sustainable development goals target of less than 12 neonatal deaths per 1000 live births (17).

A distinct trend in the decline of ANC4 visits was observed before the first cases of the COVID-19 pandemic with no sufficient evidence to justify the decline. However, the decline of ANC4 visits was noted at both KGH and WBC during the COVID-19 pandemic, although it did not reach statistical significance when compared to the periods before and after the pandemic. This finding aligns with a study in Ethiopia, which similarly found no statistical difference in the frequency of ANC4 visits during the COVID-19 pandemic compared to the pre-pandemic era(18). Despite community awareness campaigns that emphasize the importance of ANC attendance by the Ministry of Health and Partners In Health, there is still a modest impact of the COVID-19 pandemic on ANC4 visits. This impact manifested in two sharp declines following the initial reports of COVID-19 in March 2020 and a subsequent wave in December 2020. Nevertheless, ANC4 visits recovered swiftly thereafter, underscoring the resilience of the health system following public health emergencies.

Overall, there were disruptions in deliveries and ANC first visits at the national level during the COVID-19 pandemic (8). Unlike a previous study conducted in Sierra Leone (7), our study indicates that facility deliveries were relatively lower during the COVID-19 pandemic, although there was no statistically significant difference across the three periods. These low rates of deliveries during the pandemic likely reflect the mitigation strategies employed by both Partners In Health and the Ministry of Health during the pandemic to strengthen maternal health services utilization in Kono District. Robust structural and functional maternal referral systems were in place to halt a potentially major impact of the COVID-19 pandemic in this setting. Additionally, communication plans were developed to tackle misinformation campaigns, including efforts to address community members’ fear of becoming infected at health facilities.

We found that the rate of maternal mortality and neonatal mortality declined over time even during the COVID-19 pandemic. This finding is contrary to the national reports of maternal and neonatal mortality (1). The possible justification for this could be attributed to the comprehensive maternal and child health service provided by Partners In Health Sierra Leone and the heterogeneous distribution of COVID-19 cases across the country, with fewer cases in Kono district (14). Furthermore, these findings suggest resilience in maternal healthcare service due to lessons learned from the Ebola outbreak response strategies implemented in these two facilities. Our findings have important clinical and policy implications in strengthening maternal and neonatal health services in Partners In Health-supported health facilities and designing tailored strategies to improve maternal and neonatal care in the local context. These efforts are crucial for mitigating the impact of future health emergencies and ensuring continuous access to essential maternal and neonatal healthcare services.

The findings of this study should be interpreted in light of the following limitations. First, our rationale for focusing on two specific facilities was because of efforts made to support the post-Ebola recovery of these two facilities. However, this focus is affected by the small sample size of pregnant women and neonates recruited in this study. As a result, we have not been able to observe significant changes in maternal service delivery over time. Data on maternal and neonatal services from a larger number of facilities should be included in future studies. Second, the data used in our study is also limited to that available for programmatic monitoring, which does not allow us to explore variables that could provide a deeper understanding of access and uptake of maternal services. For example, we were not able to conduct longitudinal analyses at the individual level nor did we have estimates of the number of pregnancies. Therefore, we are unable to generalize the findings on ANC visits, deliveries, and maternal and neonatal outcomes to other areas of Sierra Leone.

## Conclusion

In Sierra Leone, the efforts put into place and resourced as part of the post-Ebola recovery efforts were tested during and after the COVID-19 emergency. Our study demonstrates the resilience of two healthcare facilities in a less-affected region of Sierra Leone, to the anticipated disruptions due to the COVID-19 pandemic. Further evaluation and research are recommended to sustain these achievements in future public health emergencies.

## Declaration

## Data Availability

The datasets (https://doi.org/10.5061/dryad.rxwdbrvkx) used and/or analyzed for this study are available from: http://datadryad.org/stash/share/TLn8N9UKXR9IF29JaOmw-PxDCoSjWIeAS2lQlcqkGI4.

## Acknowledgments

We acknowledge the Sierra Leone Ministry of Health and Partners In Health Sierra Leone for providing the enabling environment to conduct this study. We also want to thank Sustainable Health Systems for their partnership and collaboration in leading this effort. We thank Kristy Yiu (Unity Health Toronto) for her early support in coordinating the project. Finally, we want to express our profound gratitude to Mohamed Bailor Barrie, Jean Gregory Jerome, Kilongo Papy Mulailwa, and Pierre Ricard Pognon for their unwavering support.

## Author’s contribution

FB, ML, MMK, SL, SS, and AUN conceptualized and designed the study, with input from AKC and SM. FB and BM collated the aggregated data. ZAM and FB managed and analyzed data, with input from SM. FB and ZAM drafted the manuscript, with edits from AKC, SM, and SL. All co-authors reviewed and approved the final manuscript.

